# Actigraphic Screening for Rapid Eye Movement Sleep Behavior Disorder

**DOI:** 10.1101/19001867

**Authors:** Šandala Kristián, Dostálová Simona, Nepožitek Jiří, Ibarburu Lorenzo y Losada Veronika, Dušek Petr, Růžička Evžen, Šonka Karel, Kemlink David

## Abstract

**Background:** The patients suffering of the rapid eye movement sleep behavior disorder (RBD) are in high risk of developing a neurodegenerative disorder, most frequently from the group of alpha-synucleinopathies, such as Parkinson’s disease (PD), Dementia with Lewy Bodies (DLB) or multiple system atrophy (MSA). The definitive diagnosis of RBD is based on polysomnographic investigation. Actigraphy is much easier to perform and reflects condition in patient’s home environment.

**The aims:** The aim of this study was to find suitable biomarkers for RBD, which can be detectable by actigraphic recording.

**Methods:** High resolution actigraphic recording (MotionWatch, CamNtech ltd.) and confirming polysomnographic recording was performed on 45 RBD patients, 30 patients with other sleep-related motor disorders and 20 healthy controls. Each individual file was analysed by software testing for amount of sleep (MotionWare 1.1.20) and secondly for periodic motor activity (PLMS analysis 1.0.16). The 13-item patient self-rating RBD screening questionnaire (RBD-SQ) translated to Czech language was also used for screening purposes. We used an RBD-SQ score of five points as a positive test result, as suggested by the original publication of the scale.

**Results:** When using the actigraphic sleep detection, we encountered significant differences mostly on non-dominant hand, related to sleep fragmentation - most notably increased percentage of Short immobile bouts (47.0% vs. 28.0%, p<0.0001), increased Fragmentation index (72.5 vs. 40.7, p<0.0001) and decreased percentage of Sleep efficiency (72.1% vs. 86.8%, p<0.0001)in RBD subjects compared to other sleep disorders and controls. When analyzing periodic motor activity, we also found surprisingly more periodic hand movements (p=0.028, corrected for multiple testing), but differences on lower extremities using either measurement were not significant. The discrimination function based on RBD-SQ and Short immobile bouts % could allocate correctly the RBD status in 87.6% of cases with Wilks Lambda 0.435 and p<0.0001.

**Conclusion:** In our single-center study in patients from the Czech population, we found that actigraphic recording from upper extremities shows consistently more prominent sleep fragmentation in RBD patients compared to other sleep diagnoses or healthy controls. Actigraphy may be useful in broader screening for RBD.

## Background

REM sleep behavior disorder (RBD) belongs to the REM-related parasomnias ^1^. Its prevalence in the population aged 40-80 years is 1.06%, with the mean age of onset 61 years ^2, 3^. In contrast with male predominance observed in sleep medicine centers, both sexes are equally affected in the middle-to-older age population cohort^2^. RBD is characterized by abnormal vocalizations and/or motor activity during the REM sleep. Impaired muscle atonia during the REM sleep is a next essential feature. Motor activities may vary in intensity and complexity from short jerky limb movements to complex motor behavior and usually correlate with simultaneously experienced dream content. Nature of such dreams is mostly unpleasant, patient is often placed into position of a victim, usually attacked or being chased by unknown people or animals. Therefore, performed dream enactment can be potentially injurious to the patient or to the patient’s spouse ^1^. Previous longitudinal studies have shown that patients diagnosed with idiopathic form of RBD (IRBD) are in high risk of developing a neurodegenerative disorder from the group of alpha-synucleinopathies, e.g. Parkinson’s Disease (PD), Dementia with Lewy Bodies (DLB) and Multiple System Atrophy (MSA). Hence IRBD is considered to be a prodromal stage of underlying neurodegenerative process. Mean interval of IRBD conversion to PD has been estimated 14±6 years from the onset of the IRBD^4^. Recent prospective multicenter study by Postuma et al. has confirmed high risk of IRBD conversion to symptomatic neurodegenerative disorder and has estimated overall conversion rate 6.25% per year. Annual disease risk shows growing trend, from 10.6% after two years up to 73.5% risk of conversion after 12-year follow-up^5^. Long disease-free interval creates space for a potential neuroprotective treatment which would slow down and/or eventually stop the progressive neuronal loss ^4, 6^. For a therapeutical intervention to be possible, it is important to detect early signs of prodromal Lewy Body pathology. IRBD is the most robust clinical marker of such a disorder ^7^.

Video polysomnography (VPSG) is a current golden standard for an assessment of RBD ^1^. VPSG demands presence of trained personnel, is time-consuming, costly and not always available Highly sensitive and sufficiently specific screening tool is essential for an adequate indication of VPSG. Self-administered questionnaires are widely used as a screening method for an early detection of possible RBD. The RBD Screening Questionnaire (RBD-SQ) published in 2007 by Stiasny-Kolster et al. is very commonly used. It consists of 13 points with cut off value five points. Results above the cut off value are recommended for further clinical investigation and VPSG, eventually ^8^. Benefits of patient-administered questionnaires are simple use and interpretation of results. However, diagnostic value of screening questionnaires can be substantially lower due to possible absence of patient’s awareness of RBD symptoms namely without previous expert interview ^9-11^.

Widespread deposition of alpha-synuclein throughout structures of central, peripheral and autonomic nervous system ^12^ creates wide spectrum of symptoms, making clinically silent process of neurodegeneration detectable by various screening approaches ^13^. Besides of patient-administered questionnaires and single items primarily examining sleep-related complaints ^8, 14-17^, rating scales originally designed for evaluation of PD can also be useful in the early detection of Lewy Body pathology^13^. Mentioned rating scales, e.g. The Non-Motor Symptom Questionnaire (NMSQuest)and Movement Disorder Society-Unified Parkinson’s Disease Rating Scale (MDS-UPDRS), screen for non-motor symptoms and/or even slight motor impairment, which can be present for a long period of time before the cardinal manifestation of PD ^18, 19^. Subclinical impairment of motor function in the RBD patients can be easily examined by simple tests, e.g. Purdue Peg Board, alternate tap test and Timed ‘Up and Go’ ^20^. Nevertheless, there are more sophisticated instruments for detection of subtle motor dysfunction. Quantitative speech assessment could be enhancing component of screening, capturing articulatory motor deficits prominent in RBD subjects. Accordingly scored severity of speech impairment should correspond to significance of motor disability due to neuronal loss ^21^. Additionally, fully automated vocal evaluation which identifies and analyzes abnormalities of speech is already available ^22^. Loss of olfaction is one of the most prominent non-motor symptoms of the underlying alpha-synucleinopathy ^20^. Furthermore, olfactory dysfunction can have a substantial predictive value for the early conversion of IRBD to PD or DLB. Olfactory function can be assessed by Sniffin’ Sticks test, mainly showing impaired odor identification ^23^ and UPSIT-40, which both correlate with RBD status and UPSIT-40 scores correlated also to atrophy of grey matter in olfactory regions as measured by MRI voxel-based morphometry ^24^. The most common manifestation of autonomic dysfunction in RBD and PD subjects is constipation. Other symptoms of dysautonomia comprise cardiovascular, urinary, sexual, sudomotor and pupillomotor ^25^. Severity of autonomic dysregulation positively correlates with risk of developing an apparent alpha-synucleinopathy^26^. Patients’ report of autonomic symptoms is essential for the assessment, using NMSQuest or Scales for Outcomes in PD-Autonomic (SCOPA-AUT) ^18, 27^. Central nervous system is primarily affected, resulting in cognitive deterioration involving executive dysfunction and event-based prospective memory impairment ^28, 29^.In accordance to these findings, tests assessing attention and executive function have shown to be valuable in prediction of DLB in RBD patients ^30^.Nuclear medicine imaging techniques, such as Positron Emission Tomography (PET) and Single-Photon Emission Computed Tomography (SPECT), are able to detect even slight reduction of striatal dopamine transporter (DAT) density in RBD patients ^31, 32^. Additionally, regional alteration of cerebral metabolism can be observed using 18F-fluorodeoxyglucose PET ^33^. Measures of basal ganglia connectivity by resting-state functional magnetic resonance imaging (rs-fMRI) may also identify early state of basal ganglia disturbance ^34^. Advanced neuroimaging represents remarkably sensitive and accurate approach of RBD detection^35^, although it is not widely available and can be time-consuming. Transcranial Sonography (TCS) is accessible and easy to perform method, visualizing potential substantia nigra hyperechogenity in RBD subjects. In spite of TCS benefits, hyperechogenity of substantia nigra is found in relatively small number of RBD patients. Therefore, negative TCS has low diagnostic and prognostic value ^36^. Some of above mentioned biomarkers are used to define criteria for prodromal stage od PD ^37^ and these criteria were also validated in RBD patients, in whom they also indicated possible early conversion into Lewy-body pathology. ^38^.

Actigraphy is a promising candidate method for this purpose. The actigraphic device is wrist-watch sized triaxial accelerometer, which records bouts of motor activity. It is comfortable to wear, doesn’t alter patient’s sleeping pattern, can be used in home conditions and is suitable for long-term monitoring. Actigraphy is affordable, easy to perform and to score ^39-41^. Study performed by Louter et al. estimated specificity of 95.5% and sensitivity of 20.1% for RBD diagnosis using total wake bouts as main actigraphic discriminator [13]. A recent research by Stefani et al. found 85%-95% sensitivity and 79%-91% specificity using subjective visual expert based scoring in combination with patient self-administered questionnaires [14]. It is possible that objective, automatically calculated, quantitative actigraphic parameter in a combination with self-administered questionnaire may be sufficient yet still simple screening method for RBD in general population. The aim of this study is to find such suitable diagnostic biomarkers for RBD, which can be detectable by actigraphic recording.

## Methods

### Participants

All patients participating in the study were recruited in the Centre for disorders of sleep and wakefulness, Department of Neurology, First Faculty of Medicine, Charles University and General University Hospital, Prague.

The study was conducted in three steps

1. Discovery phase, in which the patient sample consisted of total 70consecutive subjects (mean age 60.7y, SD 13.2y, 85.7% males), comprising 20 newly diagnosed RBD patients (mean age 64.8y, SD 11.2y, 85.0% males), 30 patients diagnosed with other sleep-related motor disorders (mean age 51.5y, SD 12.0y, 83.3% males) and 20 healthy controls (mean age 70.4y, SD 6.3y, 90.0% males). Other neurological diagnoses with possible impact on sleep pattern were as follows: 1) Parkinson’s Disease (PD) with or without RBD (15 patients); 2) obstructive sleep apnea (OSA) (ten patients); 3) NREM-related parasomnias (two patients); 4) restless Legs Syndrome / Periodic Limb Movements of Sleep (RLS/PLMS) (two patients); 5) narcolepsy (one patient). Each individual included in Discovery sample underwent one night VPSG along with high resolution actigraphic recording for all four extremities, and additionally self-administered the Czech RBD-SQ^42^.
2. Replication phase – the sample consisted of 29 previously diagnosed RBD patients (mean age 69.9y, SD 7.8y, 89.6% males). RBD diagnosis was established by VPSG according to valid international criteria ^1, 43^, maximum time from the diagnosis was 3 years. Each patient included in Replication sample underwent actigraphic recording at home for multiple nights (minimal number of nights 2, maximal number of nights 6, mean number of nights 4.8, modus 6). Recording device was worn only on non-dominant hand. Participants were instructed to fill the sleep diary after every nocturnal sleep period. The items included in the sleep diary were as follows: 1) time of going to bed; 2) time of falling asleep; 3) number and duration of arousal episodes during the night; 4) time of waking up; 5) time of getting up. Patients from the Replication sample were also asked to complete the RBD-SQ.
3. Combined phase – Actigraphic and clinical data pooled from both discovery and replication phases were analyzed in order to enrich the sample by more RBD patients. This sample consisted of 45 RBD patients (mean age 66.8y, SD 9.7y, 86.6% males), 30 other sleep diagnoses and 20 healthy volunteers (same as in the Discovery phase).

### Comprehensive evaluation

Comprehensive evaluation including clinical interview and standard neurological examination was performed on each participating individual by a sleep medicine expert and neurologist. Information collected during the clinical interview comprised demographic data, character of main sleep complaints, presence of a bed partner and history of other neurological disorders.

### RBD Screening Questionnaire

To acquire additional clinical knowledge of sleep history all study participants were asked to administer Czech version of RBD Screening Questionnaire (RBD-SQ)^8, 42^. The purpose of RBD-SQ is to explore major symptomatology typical for RBD, comprising occurrence of unpleasant dreams (two items), history of dream enactment (one item), self-awareness of limb movements in sleep (one item), presence of unusual and/or harmful behaviour during sleep (five items), sleep disturbance related to dream mentation (two items) and coherent recollection of dream content (one item). Participants completed the questionnaire in the presence of a sleep medicine expert to prevent potential misinterpretations. Czech version of RBD-SQ has cut-off value of five points, as originally estimated by the questionnaire’s authors^42^. We have obtained permission to use this scale from Mapi Research Trust, Lyon, France. Internet: https://eprovide.mapi-trust.org.

### Polysomnography

Each study participant underwent one night VPSG examination at the sleep laboratory of Department of Neurology, First Faculty of Medicine and General University Hospital. Nocturnal VPSG was performed during the period of eight hours, from 22:00 to 6:00, in accordance with international standards^44^. The examination was performed using audio/video recording digitally synchronized with PSG software package (RemLogic, version 3.4.1, Embla Systems). Main parameters registered by PSG system were as follows: 1) electroencephalogram (EEG); 2) electrooculogram (EOG); 3) surface electromyogram (EMG) of bilateral mentalis muscle and the bilateral tibialis anterior muscle, 4) nasal and oral airflow and nasal pressure; 5) thoracic and abdominal respiratory efforts; 6) oxygen saturation; 7) body position and 8) electrocardiogram (ECG), as recommended by American Academy of Sleep Medicine^44^ and additionally surface EMG of the bilateral flexor digitorum superficialis muscle ^45^. All parameters registered by VPSG were visually analyzed by sleep medicine experts. Sleep pattern characteristics were scored, including arousal events, respiratory events, periodic limb movements of sleep (PLMS)or other motor activity and sleep stages ^44^. SINBAR recommendations for evaluation of RBD were used for detection and scoring of REM sleep and REM sleep without atonia (RWA)^45^.

### Actigraphy

Every participant underwent high resolution actigraphic recording (MotionWatch, CamNtech Ltd.) as another objective method for assessment of sleep pattern features. Wrist-watch sized tri-axial accelerometer was used as a recording device. Raw data consisted of time series indicating overnight distribution of motor activity bout. The data were digitally integrated and stored in 1s epochs. Companion software (MotionWareSoftWare version 1.1.20, CamNTech) was used for import, storage and consecutive analysis of actigraphic data. In all individual actigrams with user manual input of Time in bed automated software sleep analysis was performed. Selected parameters were as follows: 1) Time in bed (TIB); 2) Sleep efficiency %, characterized as a ratio of actual time spent in sleep to time spent in bed, expressed as a percentage (SE %); 3) Wake bouts, defined as the number of awakening episodes during the recorded night (WB); 4) Mobile time %, expressed as a motor activity percentage of the assumed sleep time (MT %); 5) Short immobile bouts %, equals to the number of none registered activity episodes less than or equal to one minute divided by the total of none registered activity episodes, expressed as a percentage (SIB%); 6) Mean nonzero activity epoch, characterized as the number of activity counts divided by the number of episodes of nonzero motor activity (MNAE); 7) Fragmentation index, reflects the disintegration of sleep cycle and is calculated as the sum of the MT % and the SIB % (FI). Each actigraphic file was secondly analyzed for periodic motor activity (PLMS analysis version 1.0.16), we used the main parameter PLMI (number of PLM per hour of actigraphic recording) ^46, 47^.

### Study conditions

The use of medication modifying the sleep architecture was not allowed during the evaluation period of study. However, pharmacological treatment of depression and anxiety (and RBD) was permitted (in consideration of its indication). Age under 18 years old and overnight shift work were recognized as exclusion criteria. All diagnoses were assessed in accordance with valid evaluation criteria^1^. The informed consent was signed by each participant before entering the study. The study was conducted according to the declaration of Helsinki and was approved by the Ethics Committee of General University Hospital in Prague.

### Statitical analysis

STATISTICA, data analysis software system, version 12.0. (statsoft.com) was used for statistical analysis of data. Assessment of normal distribution of data was performed using Shapiro-Wilk test, we regarded distribution as not normal if the p value was below 0.01. Data were not normally distributed, excluding SIB %. Accordingly, parametric (T-test, Pearson’s correlation analysis) or nonparametric statistics (Mann-Whitney U test) were consequently applied. Categorical data were compared using chi-squared test. Computation of basic statistics comprised of 22 independent actigrafic parameters and for this value we applied correction for multiple testing.

Comparison of clinical data was performed by ANOVA/MANOVA in specific cases with multiple measurements in the replication phase and also for the multiparametric discrimination analysis. ANCOVA was used to correct for demographic data in specific variables.

Screening test parameters for the quantitative actigraphy analysis/ in combination with RBD-SQ have been estimated, comprising sensitivity, specificity, positive predictive values and total diagnostic accuracy. The area under the ROC curve (AUC) has been determined by Receiver operating characteristics (ROC) method^48^.P-values <0.05 were recognized as statistically significant.

## Results

### Discovery phase

Intergroup analysis of actigraphic parameters found significant differences between 20 RBD patients and 50 non-RBD subjects (mean age 59.0y, SD 13.7y, 86.0% males). Age-sex structure was not significantly different between the groups. Significant differences were related to sleep fragmentation and were encountered mostly on non-dominant upper extremity. Detected between-group differences were as follows, p values after correction for multiple testing were used for interpretation: 1) increased percentage of SIB %; 2) increased Fragmentation index; 3) increased Mobile time percentage; 4) increased Wake bouts; (5) decreased Sleep efficiency. The results are summarized in Table 1. We also found surprisingly more periodic hand movements (p=0.028, corrected for multiple testing).

**Table 1.** Intergroup analysis of actigraphic parameters in Discovery phase

| Variable | Mean non-RBD | Mean RBD patients | N non-RBD | N RBD patients | Std. Dev. non-RBD | Std. Dev. RBD patients | Mann-Whitney U Test p -value |
| --- | --- | --- | --- | --- | --- | --- | --- |
| Age | 59.1 | 64.9 | 50 | 20 | 13.8 | 11.2 | 0.0655 |
| RBD-SQ | 3.4 | 8.3 | 39 | 17 | 3.1 | 3.0 | 0.0000* |
| TIB | 459.5 | 452.8 | 50 | 20 | 33.3 | 41.7 | 0.6941 |
| SE % | 86.8 | 77.4 | 50 | 20 | 8.1 | 18.6 | 0.0120 |
| WB | 35.6 | 47.8 | 50 | 20 | 15.0 | 21.4 | 0.0071 |
| MT % | 12.7 | 23.6 | 50 | 20 | 7.5 | 13.7 | 0.0005* |
| SIB % | 28.0 | 46.6 | 50 | 20 | 12.2 | 14.1 | 0.0000* |
| MNAE | 29.2 | 18.8 | 50 | 20 | 24.2 | 7.3 | 0.0224 |
| FI | 40.7 | 70.2 | 50 | 20 | 18.7 | 27.4 | 0.0000* |

\*the p-value remains significant after correction for 22 independent multiple testing.
TIB, Time in bed; SE %, Sleep efficiency %; WB, Wake bouts; MT %, Mobile time %; SIB %, Short immobile bouts %; MNAE, Mean nonzero activity epoch; FI, Fragmentation index; RBD-SQ, Rapid eye movement sleep behavior disorder screening questionnaire total score; Valid N, Valid number.

The RBD patients also significantly differed from the 20 healthy controls, but the groups did not show significant differences in age and sex structure, but there were differences in following actigraphic variables (p values are presented after correction for multiple testing): 1) increased percentage of SIB % (46.5% vs. 30.9%, p=0.0009); 2) increased Fragmentation index (70.2 vs. 45.0, p=0.0018); 3) decreased Mean nonzero activity epoch (18.8% vs. 32.6%, p<0.0001).

### Replication phase

Testing for intra-individual variability in SIB % was performed using MANOVA with replication and found no significant variability in time. Paired Sample t-Test showed no statistically significant differences in SIB % between first two consecutive nights. Power analysis of Paired Sample t-Test estimated 90% power to detect a difference lesser than one in the absolute SIB% value.

### Combined phase

Comparison of all groups combined (N=95, mean age 62.7y, SD 12.5y, 86.3% males) was performed. Significant between-group differences were as follows, p values are presented after correction for multiple testing: 1) increased percentage of SIB %; 2) increased Fragmentation index; 3) increased Mobile time percentage; 4) increased Wake bouts;5) decreased percentage of Sleep efficiency. Between-group differences in mean age also showed statistical significance. Intergroup comparison of sex structure showed no significant differences. The results are summarized in Table 2.

**Table 2.** Intergroup analysis of polysomnographic and actigraphic parameters in Combined phase

| Recording | Variable | Mean non-RBD | Mean RBD patients | N non-RBD | N RBD patients | Std. Dev. non-RBD | Std. Dev. RBD patients | Mann-Whitney U Test p-value |
| --- | --- | --- | --- | --- | --- | --- | --- | --- |
|  | Age | 59.1 | 66.8 | 50 | 45 | 13.8 | 9.8 | 0.0016 |
|  | RBD-SQI | 3.4 | 9.4 | 39 | 42 | 3.1 | 2.7 | 0.0000* |
| VPSG | TIB (min) | 460.3 | 454.2 | 47 | 19 | 32.7 | 40.2 | 0.7150 |
|  | TST (min) | 338.6 | 328.5 | 47 | 19 | 67.5 | 62.2 | 0.3107 |
|  | WASO (min) | 105.2 | 107.5 | 47 | 19 | 60.7 | 44.6 | 0.6430 |
|  | SL (min) | 16.6 | 19 | 49 | 20 | 15 | 9.6 | 0.0647 |
|  | SE % | 72 | 71.4 | 49 | 20 | 15.9 | 10.8 | 0.4823 |
|  | Awakenings | 38.9 | 38.5 | 46 | 19 | 17 | 25.6 | 0.4436 |
|  | AHI | 15.7 | 16.9 | 49 | 20 | 15.3 | 14.6 | 0.6134 |
|  | PLMI | 14.5 | 29.2 | 49 | 20 | 24.9 | 41.5 | 0.0729 |
| Leftwrist | TIB | 459.5 | 484.6 | 50 | 45 | 33.3 | 85 | 0.1436 |
|  | SE % | 86.8 | 72.2 | 50 | 45 | 8.1 | 17 | 0.0000* |
|  | WB | 35.6 | 50.9 | 50 | 45 | 15 | 21.8 | 0.0000* |
|  | MT % | 12.7 | 25.4 | 50 | 45 | 7.5 | 13.4 | 0.0000* |
|  | SIB % | 28 | 47.1 | 50 | 45 | 12.2 | 15.5 | 0.0000* |
|  | MNAE | 29.2 | 26.7 | 50 | 45 | 24.2 | 14.4 | 0.9143 |
|  | FI | 40.7 | 72.5 | 50 | 45 | 18.7 | 28.1 | 0.0000* |

\*the p-value remains significant after correction for 22 independent multiple testing.
VPSG, Video polysomnography; TIB (min), Time in bed (min); TST (min), Total sleep time (min); WASO (min), Wakefulness after sleep onset (min); SL (min), Sleep latency (min); SE %, Sleep efficiency %; AHI, Apnea-hypopnea index; PLMI, Periodic limbs movement index; TIB, Time in bed; SE %, Sleep efficiency %; WB, Wake bouts; MT %, Mobile time %; SIB %, Short
immobile bouts %; MNAE, Mean nonzero activity epoch; FI, Fragmentation index; RBD-SQ, Rapid eye movement sleep behavior disorder screening questionnaire total score; Valid N, Valid number.

Actigraphic sleep characteristics were also compared between all 45 subjects with assessed diagnosis of RBD (mean age 66.8y, SD 9.7y, 86.6% males, n=39) and 20 healthy controls (mean age 70.4y, SD 6.3y, 90.0% males, n=18). Significant intergroup differences were as follows, p values are presented after correction for multiple testing: 1) increased percentage of SIB %(47.0% vs. 30.9%, p=0.0001); 2) increased Fragmentation index (72.5 vs. 45.0, p=0.0002); 3) increased Mobile time percentage (25.4% vs. 14.0%, p=0.0005); 4) increased Wake bouts (50.8 vs. 38.9, p=0.0225); decreased percentage of Sleep efficiency (72.1% vs. 83.8%, p=0.0046). Age and sex differences between compared groups were not statistically significant. When correcting for age difference using ANCOVA, we still observed highly significant differences for percentage of SIB% between RBD and non-RBD subjects (p<0.0003).

In our study, RBD-SQ could allocate RBD status with 95.2% sensitivity and 71.7% specificity with cut-off of 5 points. Actigraphic detection of RBD using only SIB % as a main discriminator, had sensitivity and specificity of 86.6%and 66.0%(for cut-off 31%), 84.4% and 80.0% (for cut-off 35%), respectively. Diagnostic accuracy based on combination of actigraphy (expressed by SIB%) and RBD-SQ (including only subjects with RBD-SQ>4) could identify RBD with estimated sensitivity of 90.0% (for cut-off 31%) / 87.5%(for cut-off 35%) and specificity of 81.8%, respectively. AUC of the combined screening method was 0.845.When filtering all subjects for the SIB %> 31%, the sensitivity and specificity of subsequent use of RBD-SQ is 100% and 83.3%. When using both cut-offs of SIB %>31 and RBD-SQ>4 for positivity of screening and all other cases were regarded as negative, the sensitivity and specificity was85.7% and 97.4%, respectively. Comparative results of all ROC analysis including confidence intervals are presented in table 3. Complete results of ROC analysis are provided in the Supplementary material.

**Table 3.**
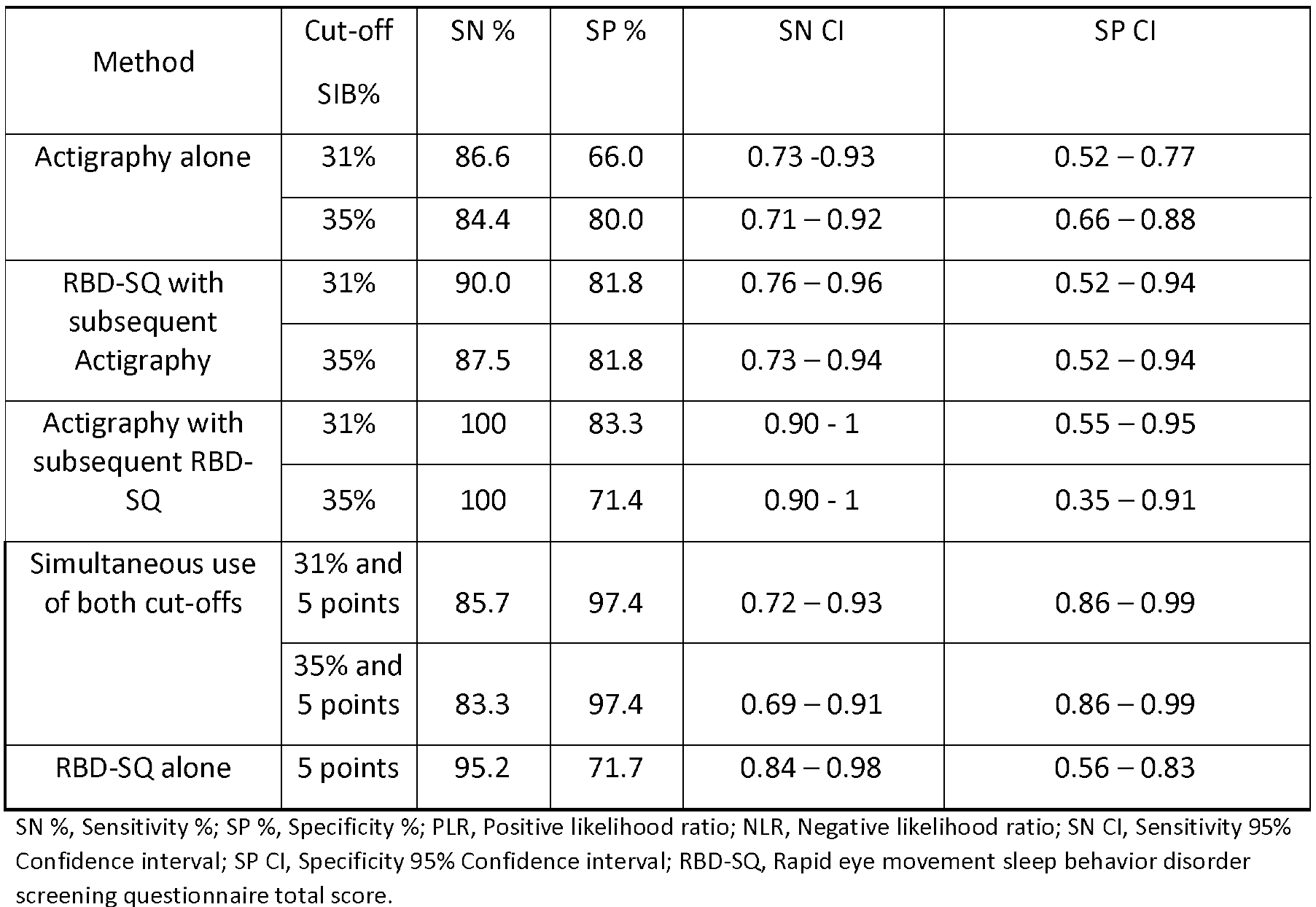
Comparative selected results of ROC analysis

We performed stepwise discrimination analysis using all parameters significantly different between RBD and non-RBD subjects. In the first step we had to exclude Wake bouts and Mobile time percentage for their high correlation with Sleep efficiency %. All parameters with F lower than two were subsequently removed. In the end of the analysis RBD-SQ and SIB % were the only remaining parameters in the model.

The discrimination function based on RBD-SQ and SIB % could correctly allocate the RBD status in 87.6% of cases with Wilks Lambda 0.435 and p<0.0001.

### Implication of ROC results in the Replication phase

There are eight of total 29 actigraphic recordings from the Replication sample which had multiple night recoding and showed at least one night with SIB %below the cut off value of 31%. Out of those eight cases, in two RBD patients the SIB % values during all recorded nights were below the cut off value of 31%. In six patients, at least one of the recorded nights had SIB % value above 31%.

## Discussion

Our study fulfilled its aim, which was to determine robust biomarkers of RBD detectable by actigraphy. We demonstrated the effectiveness of combination of RBD-SQ and actigraphicaly assessed sleep fragmentation - in our study best represented by SIB%.

Patient’s perception of sleep disturbance is an essential feature in the current diagnostics of RBD. Nevertheless, 44% of patients suffering of RBD are not aware of occurring dream-enactment behaviour. This may be due to absence of the bed partner or due to unimpaired quality of sleep in the 70% of RBD patients ^11^. Patient’s lack of self-awareness of RBD symptoms leads to not seeking medical attention and makes potential therapeutic intervention impossible. Therefore, development of unbiased and widely available screening method for RBD in general population is of high importance. Additionally, it is relevant to avoid false negative screening results and to reduce the number of eventually negative VPSG, at the same time.

In our study, conducted analysis of actigraphs showed notable differences in selected actigraphic parameters between groups throughout all phases of the study. The intergroup differences were sufficiently significant to distinguish patients with assessed diagnosis of RBD from the other study participants, comprising patients with other sleep-related motor disorders and healthy controls. The most prominent differences were encountered on non-dominant upper extremity and were highly related to increased sleep fragmentation in the RBD patients. In accordance with the results of statistical analysis, we determined SIB %as a best actigraphic discriminator of RBD status. SIB %is stable when comparing patient’s home vs. sleep laboratory recording and is an equivalent to Short burst inactivity index^41^.

Previous studies made significant progress in evaluating the actigraphy as a screening tool. Findings of Louter et al. showed that using Total wake bouts as a main actigraphic discriminator could identify RBD only in 20.1% of the cases. Such sensitivity is not sufficient for the clinical screening ^40^. More recent study of Stefani et al. used subjective expert-based analysis of actigraphic record, visually scoring occurrence and amplitude of wrist activity during the nocturnal sleep period. Raters were provided basic clinical data and actigraphs were subsequently compared with VPSG for association with selected polysomnoghraphic parameters. Visual actigraphy analysis in combination with patient-administered questionnaires had estimated 85%-95% sensitivity and 79%-91% specificity ^41^. This proposed combination of methods shows good overall sensitivity and specificity at the expense of easy scoring, potentially. Visual scoring of long term actigraphic record would possibly be time-consuming and would require well-trained sleep medicine expert. Despite its other qualities, these features of the visual scoring would practically limit its usage as a screening in the general population, e.g. in the fully self-administered screening process. Additionally, interpretation of both used methods highly depends on a human factor – patient’s awareness of symptoms and expert’s rating skills. When using objective quantitative actigraphic analysis separately of screening questionnaires, Stefani et al. found significantly different Short burst inactivity index between RBD patients and healthy controls, but not between RBD patients and other sleep-related motor disorders. This led to conclusion that quantitative actigraphy is not a sufficient screening method for RBD ^41^.

According to our findings, single-wrist actigraphy using SIB % could identify RBD cases among other sleep-related diagnoses and healthy controls with estimated sensitivity and specificity of 86.6% and 66.0% (for cut-off 31%), 84.4% and 80.0% (for cut-off 35%), respectively.

In our study, RBD-SQ as a routinely used screening instrument, could allocate RBD status with higher sensitivity of 95.2%, but with lower specificity of 71.7%, when compared to SIB %.When testing both methods combined, we used two different RBD screening approaches: 1)using positive result of RBD-SQ (cut off 5 points) as a criterion for subsequent actigraphy recording and 2) using positive result of actigraphy as a criterion for following RBD-SQ administration. Further research consequently showed that both combinations of methods have preferable screening test characteristics. When usingRBD-SQ followed by actigraphic recording, we estimated 90.0%sensitivityfor cut-off value of 31% and 87.5% sensitivity for cut-off value of 35%, respectively. Specificity of 81.8% increased and was equal in both cut-off values. When using actigraphy first with subsequent administration of RBD-SQ, we estimated sensitivity and specificity of 100% and 83.3% (for cut off 31%), 100% and 71.4% (for cut off 35%), respectively. Our study showed that conjunction of quantitative actigraphy analysis and RBD-SQhas sufficiently high sensitivity and promotes specificity of screening for RBD. We suggest the bestSIB % cut-off value of 31%, when combined with the positive result of RBD-SQ.

There are possible study limitations which must be noted. Majority of the study participants was represented by the males. In spite of this fact, sex ratio was similar in all groups and showed no significant intergroup differences. Another possible limitation of the study was that patients with confirmed diagnosis of RBD were significantly older than other study participants, comprising controls and patients with other sleep-related disorders. These study sample characteristics correspond to typical clinical features of the RBD, as the vast majority of diagnosed patients are men of the older age^3^, however, the inter-group differences in SIB % remained significant after correction for age as a possible confounder.

Some of our RBD subjects were under antidepressants treatment. Even though drug intake may trigger manifestation of latent pre-existing RBD, we suggest that the motor patterns of RBD with and without antidepressants are similar^1, 49^. Moreover, in accordance to our current opinion, psychoactive medication should not be able to induce the deposition of alpha-synuclein de novo.

Potential strengths of our study were as follows: 1) relatively high number of patients diagnosed with various sleep-related motor disorders; 2) all study participants, including healthy controls, underwent standardized VPSG examination; 3) all participants included in the Discovery phase underwent VPSG along with actigraphic recording of all four extremities, accordingly allowing comparison of actigraphic record with simultaneously detected EMG activity.

We would like to point out potential extended use of actigraphy in the future diagnostic process. It is possible that pooled actigraphic recordings from various centers might be suitable for machine learning approaches. Compact design and comfortable use could make actigraphy an ideal screening tool in the general population, not only in the population in a high risk of RBD. Moreover, existing smartphone gadgets are able to record similar type of data as actigraphs. Every person is in a potential risk of developing a neurodegenerative disorder and people above 60 are at higher risk, as the mean age of RBD onset is 61 years ^3^. Broad implementation of actigraphy (either through detection of high SIB%, or future more advanced machine learning approaches) could represent the first step of automatically delivered RBD screening program, self-administered in patient’s home conditions, which in next steps could include also RBD-SQ and speech testing^21^, whereas other screening procedures (in depth described in background) are either costly and/or time consuming. Subsequent positivity of this comprehensive at home screening method would lead to suggestion to the user for an appointment with the local sleep medicine centre for further medical investigation including VPSG, eventually. Such simple screening strategy could provide early detection of emerging alpha-synucleinopathy in population of smart phone users with a watch gadget and thus make potential neuroprotective treatment available for broader, unaware and unselected group of people at risk.

## Conclusion

We proved our research hypothesis that automatic quantitative analysis of actigraphic recoding in combination with RBD-SQ is accurate, highly sensitive and specific method for the detection of RBD. We suggest that quantitative actigraphy and RBD-SQ combined provide balanced ratio of required sensitivity and specificity. Therefore, we suggest introduction of single-wrist actigraphic recording into the RBD screening algorithm.

## Data Availability

The datasets used and analysed during the current study are available from the corresponding author upon reasonable request

## Acknowledgements

The authors greatly appreciate all subjects who volunteered to participate in the experiments described in this paper.

The permission to use the RBDSQ was obtained from Mapi Research Trust, Lyon, France. Internet: https://eprovide.mapi-trust.org.

